# Plasma Small RNA profiling reveals a predictive three-miRNAs signature for early beta cell dysfunction across glucose tolerance stages

**DOI:** 10.64898/2025.12.02.25341443

**Authors:** Elena Aiello, Giuseppina Emanuela Grieco, Michela Brunetti, Giulia Gliozzo, Daniela Fignani, Giuseppe Quero, Sergio Alfieri, Gianfranco Di Giuseppe, Gea Ciccarelli, Angela Dardano, Martina Parenti, Andrea Mari, Roberto Bizzotto, Giuseppe Daniele, Andrea Giaccari, Francesco Dotta, Guido Sebastiani, Teresa Mezza

**Affiliations:** Diabetes Unit, Department of Medicine, Surgery and Neurosciences, University of Siena, Siena-Italy; Fondazione Umberto Di Mario ONLUS c/o Toscana Life Science, Siena-Italy; Tuscany Centre for Precision Medicine (CReMeP), Siena-Italy; Dipartimento di Medicina e Chirurgia Traslazionale, Università Cattolica del Sacro Cuore, Rome, Italy; Endocrinology, Diabetes and Internal Medicine, Fondazione Policlinico Universitario Gemelli IRCCS, Roma, Italy; Chirurgia Digestiva, Fondazione Policlinico Universitario Agostino Gemelli IRCCS, Roma, Italy; Department of Clinical and Experimental Medicine, University of Pisa, Pisa, Italy; Institute of Neuroscience, National Research Council, Padova, Italy

**Keywords:** Beta cell function, microRNA, Prediction, Type 2 Diabetes

## Abstract

**Aims/hypothesis:** Type 2 diabetes mellitus (T2D) is a multifactorial disease marked by progressive beta cell dysfunction; however, no reliable, easily measurable and non-invasive circulating biomarkers are currently available to track this decline. Since microRNAs (miRNAs) tightly regulate beta cell physiology and are secreted and found in blood, we investigated whether specific circulating miRNAs may reflect in vivo beta cell function in living donors.

**Methods:** We conducted a cross-sectional study on two independent cohorts: a discovery cohort composed by 78 individuals with different glucose tolerance [23 with normal glucose tolerance (NGT), 22 with impaired glucose tolerance (IGT), and 33 with T2D] and a validation cohort composed by 158 subjects [71 non-diabetic (ND) and 87 with T2D] who were administered an oral glucose tolerance test (OGTT) and/or a mixed meal test (MMT) with measurement of glucose, insulin and C-peptide. Plasma RNA was profiled by small RNA sequencing and differences between glucose tolerance groups were analysed using DESeq2. Linear regression analyses were used to associate miRNA abundance to clinical and metabolic measures. Droplet Digital PCR (ddPCR) was used to validate selected miRNAs. A LASSO model was used to select a set of features predictive of beta cell function.

**Results:** We identified eleven differentially expressed miRNAs across the three glucose tolerance groups of the discovery cohort. Integrated analyses pinpointed a three-miRNAs signature (miR-34a-5p, miR-1306-5p and miR-335-5p) each significantly correlated with beta cell rate sensitivity (RS), an early indicator of insulin secretory impairment. Incorporating this panel with age and 1-hour post-load glucose into a LASSO regression model enabled accurate prediction of RS (Spearman ρ=0.43, p<0.05). Model performance was confirmed in the validation cohort of 71 ND and 87 T2D subjects (Spearman ρ=0.23, p<0.05). To streamline its clinical use, we substituted fasting glucose for 1-hour post-load glucose while retaining miRNA levels and age; this adaptation preserved predictive power (Spearman ρ=0.46, p<0.05 discovery; ρ=0.15, p=0.06 validation).

**Conclusion/interpretation:** This three-miRNAs signature combined with simple clinical parameters constitutes a promising, non-invasive biomarker for early beta cell dysfunction and T2D progression.

**Research in context:** *What is already known about this subject?:*

- Beta cell dysfunction is a key event in Type 2 Diabetes (T2D) pathogenesis
- Circulating microRNAs (miRNAs) are biomarkers linked to T2D onset and T2D-associated metabolic abnormalities

*What is the key question?:*

- Can specific circulating miRNAs serve as reliable biomarkers of beta cell dysfunction across different stages of glucose tolerance and Type 2 Diabetes (T2D)?

*What are the new findings?:*

- Small RNA profiling of plasma samples from deep metabolically characterized NGT, IGT, and T2D subjects revealed circulating miRNAs significantly associated with in-vivo beta cell function, reflecting key mechanisms underlying pancreatic beta cell physiology
- A specific set of three circulating miRNAs- miR-34a-5p, miR-1306-5p and miR-335-5p - resulted linked to beta cell dysfunction
- This three-miRNA signature can predict early alterations in insulin secretion, representing a promising non-invasive biomarker for the early identification of individuals at risk of developing T2D

**How might this impact on clinical practice in the foreseeable future?:** Integrating circulating miRNA profiling with routine baseline clinical parameters could provide a simpler, non-invasive, and more accessible method to identify individuals at risk of beta cell dysfunction and T2D. Implementing miRNA-based diagnostics could facilitate personalized monitoring of disease progression.

**Graphical Abstract:** 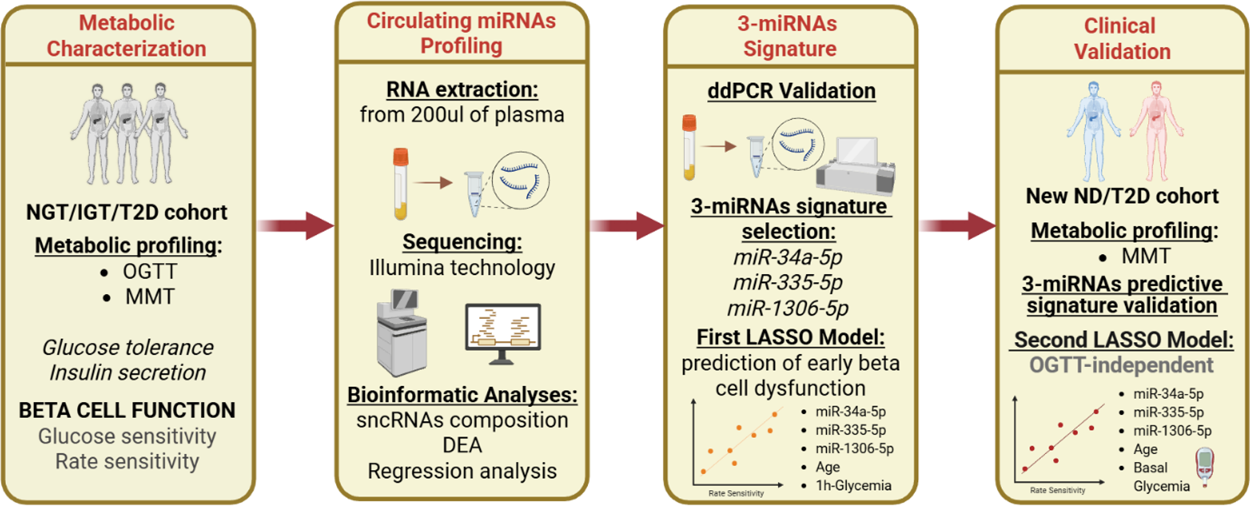

## Introduction

Type 2 Diabetes (T2D) is a chronic metabolic disorder characterized by a progressive decline in functional insulin-producing pancreatic beta cells and heightened peripheral insulin resistance, leading to persistent hyperglycaemia and an increased risk of cardiovascular complications [1]. In T2D, both beta cell dysfunction [2] and the reduction of beta cell mass [3] are key contributors to the development of hyperglycaemia, particularly in the context of diminished peripheral insulin sensitivity. Our recent findings highlight that early impairments in first-phase insulin secretion and glucose sensitivity, two critical indicators of beta cell functionality, can predict the onset of hyperglycaemia [4].

These findings emphasize that functional alterations in beta cells are central to the pathogenesis of T2D. Consequently, there is a pressing need for the identification and clinical implementation of additional biomarkers that reflect the functional status of beta cells. These biomarkers could enhance our understanding of the disease and improve clinical outcomes through more tailored therapeutic strategies.

Circulating nucleic acids are considered promising clinical biomarkers. In particular, circulating microRNAs (miRNAs), short 19-22 nucleotide-long non-coding RNAs known to play a crucial role in the regulation of gene expression [5] [6], have been implicated in various diseases. In the context of T2D, numerous studies have reported alterations in the levels of specific miRNAs in multiple tissues, including insulin-sensitive tissues [7, 8], pancreatic islets and beta cells [9, 10], as well as plasma or serum of T2D affected individuals. These alterations are not only associated with the progression of T2D but are also linked to several metabolic abnormalities characteristic of the disease [11–19]. Notably, a specific set of 10 circulating miRNAs has been consistently associated with T2D [20]. These miRNAs exhibit altered expression, either increased or decreased, when compared to non-diabetic individuals, and their levels have been correlated with key clinical characteristics of T2D, such as blood glucose levels and insulin sensitivity. However, it is important to note that none of these studies directly assessed miRNAs in relation to high-quality measures of beta cell function, such as rate sensitivity (RS). Importantly, RS emerges as a more sensitive indicator of early beta cell dysfunction respect to other metabolic measures [4]; as a matter of fact, RS is significantly reduced in those individuals developing T2D after beta cell mass reduction (upon partial pancreatectomy), thus supporting the use of RS as an early functional and clinically relevant parameter to predict the onset of secretory deficits before the onset of overt T2D. Consequently, the lack of indicative and accessible biomarkers highlights the need for future research to explore the potential connection between circulating miRNAs expression and beta cell functionality, which could offer deeper insights into the pathophysiology of T2D and, more importantly, novel clinically useful biomarkers.

In this study, we linked in vivo metabolic characteristics to circulating profiles of miRNAs in the plasma of individuals with normal glucose tolerance (NGT), impaired glucose tolerance (IGT), and Type 2 Diabetes (T2D). By leveraging comprehensive metabolic profiles, including detailed mathematical modelling of beta cell functional parameters, our goal is to identify a circulating miRNAs signature that can be used as a biomarker directly associated with beta cell functionality, which may serve as a predictor for individualized prognostic evaluation and therapeutic responses in T2D.

## Materials and Methods

### Study population and metabolic profile

Seventy-eight subjects (40 males; 38 females; mean age 66.6±10.7 [years ± SD]) undergoing pylorus-preserving pancreatoduodenectomy were recruited from January 2017 to July 2019 at the Digestive Surgery Unit and studied at the Centre for Endocrine and Metabolic Diseases unit (Agostino Gemelli University Hospital, Rome, Italy). Patients were metabolically profiled before undergoing surgery. Based on the thresholds set by the American Diabetes Association (ADA), subjects were classified in normal glucose tolerant (NGT, N = 23), impaired glucose tolerant (IGT, N = 22) and type 2 diabetic (T2D) with disease onset longer than 2 years (N = 33) (**ESM Methods**). All 78 subjects underwent an oral glucose tolerance test (OGTT) and/or a mixed meal test (MMT) to evaluate functional parameters describing insulin secretion (**Table 1**; **ESM Methods**).

**Table 1.** Demographic and OGTT-clinical/metabolic characteristics of the discovery cohort. Data are reported as mean ± SD, sex distribution is presented as counts. Comparisons between groups were performed using a Kruskal-Wallis test or Ordinary ONE-Way ANOVA test according to the estimated normality of the residuals. Statistical difference for the variable ‘Sex’ was evaluated with Fisher’s exact test. P-values from pairwise comparisons were adjusted for multiple tests (Dunn’s or Tukey’s post hoc test respectively).

| Variable | NGT<br>(n=23) | IGT<br>(n=22) | T2D<br>(n=33) | p-value | Post- hoc p-value |
| --- | --- | --- | --- | --- | --- |
| Age | 62.5 ± 12.2 | 67 ± 11.1 | 69.2 ± 8.6 | 0,12 | 0,04* |
| Sex (M/F) | 10/13 | 12/10 | 18/15 | 0,77 | - |
| BMI (kg/m<sup>2</sup>) | 25.3 ± 3.7 | 25.8 ± 5.1 | 25.1 ± 4.3 | 0,74 | ns |
| HbA1c (mmol/mol) | 34.8 ± 5.81 | 42 ± 8.47 | 44.3 ± 11.4 | 0,02 | 0,03*; 0,05<sup>†</sup> |
| HbA1c (%) | 5.34 ± 0.53 | 5.99 ± 0.77 | 6.20 ± 1.05 | 0,02 | 0,03*; 0,05<sup>†</sup> |
| Basal Insulin (pmol/L) | 41.4 ± 18.3 | 50.7 ± 25.4 | 59.8 ± 67.4 | 0,64 | ns |
| Mean Insulin (pmol/L) | 290.5 ± 138 | 349.6 ± 183.9 | 210.5 ± 144.4 | 0,02 | 0,02<sup>‡</sup> |
| Basal Glucose (mmol/L) | 5.2 ± 0.6 | 5.3 ± 0.6 | 6.8 ± 1.7 | <0,01 | 0,0006*; 0,002<sup>‡</sup> |
| Mean Glucose (mmol/L) | 7.8 ± 1.4 | 9.3 ± 1.5 | 13.5 ± 3.1 | <0,01 | 0,0001*; 0,0001<sup>‡</sup> |
| 1h Glucose (mmol/L) | 9.1 ± 2.5 | 10.5 ± 2.3 | 14.6 ± 3.8 | <0,01 | 0,0001*; 0,0001<sup>‡</sup> |
| Basal Insulin Secretion Rate<br>(pmol min<sup>-1</sup> m<sup>-2</sup>) | 61.8 ± 21.9 | 78.9 ± 43 | 80.8 ± 39.2 | 0,12 | ns |
| Total Insulin Output (nmol m<sup>-2</sup>) | 41.5 ± 16.8 | 47.5 ± 17.8 | 34.2 ± 18.8 | 0,046 | 0,04<sup>†</sup> |
| Rate Sensitivity<br>(pmol m<sup>-2</sup> mM<sup>-1</sup>) | 817.5 ± 1026.4 | 441.9 ± 610.2 | 146.6 ± 252.2 | 0,006 | 0,004* |
| Glucose Sensitivity<br>(pmol min<sup>-1</sup> m<sup>-2</sup> mM<sup>-1</sup>) | 82.8 ± 34.4 | 76.6 ± 34.4 | 33.4 ± 26.4 | <0,01 | 0,0001*; 0,0002<sup>‡</sup> |
| Potential Factor Ratio | 2.1 ± 0.9 | 1.6 ± 0.5 | 1.2 ± 0.5 | <0,01 | 0,0001*; 0,01<sup>‡</sup> |

\*NGT vs T2D; <sup>†</sup>NGT vs IGT; <sup>‡</sup>IGT vs T2D
For all clinical parameters (Age, Sex, BMI, HbA1c) comparisons were performed on the total number of participants (n=23 NGT, n=22 IGT and n=32 T2D).
For *Basal Insulin*, *Mean Insulin*, *Basal Glucose*, *Mean Glucose*, *1h-Glucose*, *Basal Insulin Secretion Rate*, *Total Insulin Output*, *Glucose Sensitivity* and *Potential Factor Ratio*, comparisons were performed using OGTT-derived data for n=21 NGT, n=20 IGT, and n=27 T2D. *Rate sensitivity* was calculated using data from both the OGTT and the MMT, with comparisons based on n=23 NGT, n=22 IGT, and n=32 T2D.

In addition, to validate our findings, we enrolled 158 subjects without pancreatic diseases, who attended the Centre for Endocrine and Metabolic Diseases unit (Agostino Gemelli University Hospital, Rome, Italy) and the Diabetes outpatients clinic (Azienda Ospedaliero-Universitaria Pisana, Pisa, Italy) between July 2022 and May 2024. The subjects were divided into 71 non-diabetic controls, ND, (39 females, 32 males) and 87 patients with established T2D (29 females, 58 males), who underwent MMT and calculation of functional parameters describing insulin secretion (**Table 2**; **ESM Methods**).

**Table 2.** Demographic and MMT-derived clinical/metabolic characteristics of the validation cohort. Data are presented as mean (± SD), sex distribution is presented as counts. Comparisons between groups were performed using the Mann–Whitney U test for continuous variables. Statistical difference for the variable ‘Sex’ was evaluated with Fisher’s exact test.

| Variable | ND (n=71) | T2D (n=87) | p value |
| --- | --- | --- | --- |
| Age | 41.96 ± 13.47 | 60.47 ± 10.21 | < 0,001 |
| Sex (M/F) | 32/39 | 58/29 | <0,05 |
| BMI (kg/m<sup>2</sup>) | 26.23 ± 4.90 | 30.12 ± 5.11 | < 0,001 |
| HbA1c (mmol/mol) | 35.3 ± 4.42 | 55 ± 9.46 | < 0,001 |
| HbA1c (%) | 5.38 ± 0.41 | 7.18 ± 0.87 | < 0,001 |
| Basal Insulin (pmol/L) | 42.32 ± 32.31 | 79.91 ± 113.25 | < 0,001 |
| Mean Insulin (pmol/L) | 227.81 ± 170.88 | 344.05 ± 283.96 | 0,01 |
| Basal Glucose (mmol/L) | 5.56 ± 0.70 | 8.24 ± 2.06 | < 0,001 |
| Mean Glucose (mmol/L) | 6.27 ± 1.21 | 11.18 ± 2.92 | < 0,001 |
| 1h Glucose (mmol/L) | 6.2 ± 1.6 | 11.1 ± 3.1 | <0,01 |
| Basal Insulin Secretion Rate (pmol min<sup>-1</sup> m<sup>-2</sup>) | 60.53 ± 32.30 | 104.94 ± 63.59 | < 0,001 |
| Total Insulin Output (nmol m<sup>-2</sup>) | 47.92 ± 22.50 | 69.22 ± 32.68 | < 0,001 |
| Rate Sensitivity (pmol m<sup>-2</sup> mM<sup>-1</sup>) | 1207.83 ± 1391.13 | 454.05 ± 628.71 | < 0,001 |
| Glucose Sensitivity (pmol min<sup>-1</sup> m<sup>-2</sup> mM<sup>-1</sup>) | 97.13 ± 66.70 | 61.06 ± 65.86 | < 0,001 |
| Potential Factor Ratio | 1.44 ± 0.70 | 1.27 ± 0.61 | 0,02 |

The study protocol (ClinicalTrials.gov NCT02175459) was approved by the Ethical Committee Fondazione Policlinico Universitario Agostino Gemelli IRCCS – Università Cattolica del Sacro Cuore (P/656/CE2010 and 22573/14), and all participants provided written informed consent, followed by a comprehensive medical evaluation.

### Oral Glucose Tolerance Tests (OGTT) and Mixed Meal Tests (MMT)

A standard 75 g OGTT was performed with measurement of glucose, insulin and C-peptide at 0, 30, 60, 90, 120 min after glucose load. Based on the pre-surgery OGTT results, we classified the patients according to the ADA classification (American Diabetes Association, 2019) (see **ESM Methods**). A mixed meal test (MMT) was performed as previously described [21] (see **ESM Methods**).

In the OGTT and MMT, insulin secretion [basal insulin secretion rate (bISR) and total insulin output (tISR)] and the related functional parameters were derived from C-peptide levels via mathematical modelling: beta cell glucose sensitivity (GS) is the slope of the static relationship between insulin secretion and glucose concentration; beta cell rate sensitivity (RS) reflects early phase insulin release; potentiation factor ratio (PFR1) reflects the enhancement of insulin secretion over time, due to multiple mechanisms such as sustained hyperglycaemia, non-glucose substrates, incretin effect and neural influences [22–24] (see **ESM Methods**). For the discovery cohort, all parameters were primarily calculated from OGTT measurements. When RS was predicted via a LASSO model (see Statistical Analysis section), values derived from MMT were used in those subjects for whom OGTT data were not available.

### Blood samples collection, plasma samples processing and total RNA extraction

Blood samples were collected following a specific Standard Operating Procedure (SOP) as previously reported [25, 26] (see **ESM Methods**).

Total RNA extraction was performed from 200 µL of plasma through Serum/Plasma Norgen kit (cat. 55000, Thorold, ON L2V 4Y6, Canada) following manufacturer’s recommendation.

### Small RNA cDNA library preparation and QC

Small RNA-derived cDNA libraries were prepared using QiaSeq miRNA library kit (cat. 331505, Qiagen) as previously described [25, 26] (see **ESM Methods**). Then, libraries quality control (QC) was performed through QUBIT 3.0 spectrofluorometer (Qubit™ dsDNA HS Assay Kit, cat. Q32854, Thermofisher Scientific) and Bioanalyzer 2100 (Agilent High Sensitivity DNA kit cat. 5067-4626, Thermofisher Scientific). Following QC, all libraries were normalized and further sequenced on Illumina NovaSeq 6000 platform [NovaSeq 6000 SP Reagent Kit (100 cycles) cat. 20027464, NovaSeq XP 2-Lane Kit cat. 20021664, Illumina] using the XP protocol applying 75×1 single reads (see **ESM Methods** for details). Data were returned from BaseSpace Sequence Hub as demultiplexed FASTQ files.

### Small RNAs quantification and profiling

FastQ files from small RNA sequencing were processed using sRNAbench. Reads were processed with the Qiagen (with UMIs) protocol for adapters and duplicates removal. Reads were aligned to the human reference genome (GRCh38.p13) using Bowtie (seed = 20, ≤1 mismatch, minimum read length = 15 nt, minimum read count = 2, ≤10 multiple alignments). miRNAs were annotated using miRBase v22.1, while other small RNAs were annotated using RNAcentral v20.0 and Ensembl ncRNA/cDNA (release 104). Raw miRNA counts were extracted from the mature_sense.grouped output, and biotype distribution was derived from mappingStat files (see **ESM Methods** for details).

### Selection of miRNAs of interest for the validation stage

To select the most relevant circulating microRNAs to be included in the validation stage, we adopted the following criteria: (*i*) differentially expressed miRNAs across glucose tolerance groups with a median UMI (Unique Molecular Identifier) count from sequencing of 100 or more, and/or (*ii*) miRNAs with a median UMI count of 100 or more with almost one significant association with GS and/or RS as parameters of beta cell function.

### Reverse Transcription and ddPCR

Validation of miRNAs selected through both differential expression analysis and regression analysis was performed through miRCURY LNA Reverse Transcription and subsequent droplet digital PCR (ddPCR) detection (see **ESM Methods**).

Thresholds to separate positive from negative droplets were set manually for each miRNA using the histogram function and reads were analysed using QuantaSoft™ Analysis Pro software (Version 1.2, BioRad, Mississauga, ON, Canada).

### Statistical Analysis

In both cohorts (discovery and validation), baseline clinical characteristics were compared across glucose tolerance groups. Firstly, normality of residuals was assessed with the Shapiro–Wilk test. Depending on distribution, ANOVA or Kruskal–Wallis tests evaluated overall group differences, followed by pairwise Dunn’s or Tukey’s test (see **ESM Methods**). In the discovery cohort, age resulted significantly higher in T2D vs NGT after post-hoc comparison (**Table 1**), then included as covariate in the subsequent analyses.

Longitudinal responses during the Mixed Meal Test (MMT) and Oral Glucose Tolerance Test (OGTT) were evaluated using generalized linear mixed-effects models with Gamma distribution and log link, including fixed effects for time and glucose tolerance group and a random subject intercept. Type III Wald chi-square tests assessed main and interaction effects, and mean concentrations ± SEM were visualized over time (see **ESM Methods**).

For small RNA sequencing data, samples with less than 1×10^6^ miRNA raw reads were excluded from further analyses. Low-abundance filtering retained miRNAs with >5 CPM in at least 50% of samples, yielding 246 expressed miRNAs. Filtered miRNA counts were normalized using DESeq2 median-of-ratios, and differential expression analysis across glucose tolerance groups was performed using the Wald test, with the age as covariate. Multiple-testing correction was applied using the Benjamini–Hochberg method; miRNAs with padj<0.05 were considered differentially expressed (see **ESM Methods**).

Differential expression results were validated by droplet digital PCR (ddPCR). Log2-transformed ddPCR copies (with pseudocount) were analysed using linear models including group and age as predictors. miRNAs with statistically significant group effect (*p*<0.05) were considered confirmed (see **ESM Methods**).

Associations between miRNA expression and clinical variables were examined using linear models. Log2-transformed miRNA expression values were analysed by linear regression including age as covariates. Influential values were removed using Cook’s distance. Significant associations were defined at p<0.05, and partial R^2^ statistics were calculated. For visualization, miRNA expression was adjusted for covariate effects (see **ESM Methods** for details).

A LASSO regression model was developed to predict rate sensitivity using selected miRNAs (miR-34a-5p, miR-1306-5p, miR-335-5p) and clinical covariates. The model was tuned using leave-one-out cross-validation, and performance was assessed by correlation between predicted and observed values (see **ESM Methods**).

## Results

### Small RNA sequencing analysis of plasma samples from individuals with normal, impaired glucose tolerance and Type 2 Diabetes

A total of 78 participants (**Figure 1A**) underwent deep metabolic phenotyping, including OGTT and MMT, to quantify key aspects of glucose homeostasis, including glucose tolerance, GS and insulin secretory dynamics (i.e., RS). Subjects were stratified into three groups: n=23 with normal glucose tolerance (NGT), n=22 with impaired glucose tolerance (IGT), and n=33 with established Type 2 diabetes (T2D) (**Table 1**; **Supplementary Figure 1**). As we can see from MMT curves, individuals with T2D showed markedly higher glucose concentrations (**Figure 1B**; p < 0.001) and significantly lower insulin (**Figure 1C**; p < 0.001) and C-peptide (**Figure 1D**; p < 0.001) responses compared with NGT and IGT. As expected, mean fasting and post-load glucose progressively increased across the groups: NGT (7.7 ± 1.4 mmol/L), IGT (9.2 ± 1.4 mmol/L), and T2D (13.3 ± 3.4 mmol/L; p < 0.01 vs. NGT and IGT). Beta cell function progressively declined across tolerance stages: in particular, RS fell from 817.5 ± 1026.4 pmol m^-2^ mM^-1^ in NGT to 146.6 ± 252.2 pmol m^-2^ mM^-1^ in T2D (p< 0.01), GS decreased from 82.8 ± 34.4 pmol min^-1^ m^-2^ mM^-1^ in NGT to 33.4 ± 26.4 pmol min^-1^ m^-2^ mM^-1^ in T2D (p< 0.01), and PFR1 was reduced in T2D (1.2 ± 0.5) vs. NGT (2.1 ± 0.9; p < 0.01). (**Table 1**). tISR was also reduced in T2D compared with NGT and IGT (p = 0.04). Collectively, these data confirm a progressive deterioration in beta cell responsiveness and glucose regulation from NGT through IGT to overt T2D (**Table 1**, **Figure 1B, C, D and Supplementary Figure 1**). Small RNA sequencing analysis was conducted on plasma samples of n=78 subjects, subdivided into n=23 NGT, n=22 IGT, and n=33 T2D.

**Figure 1.**
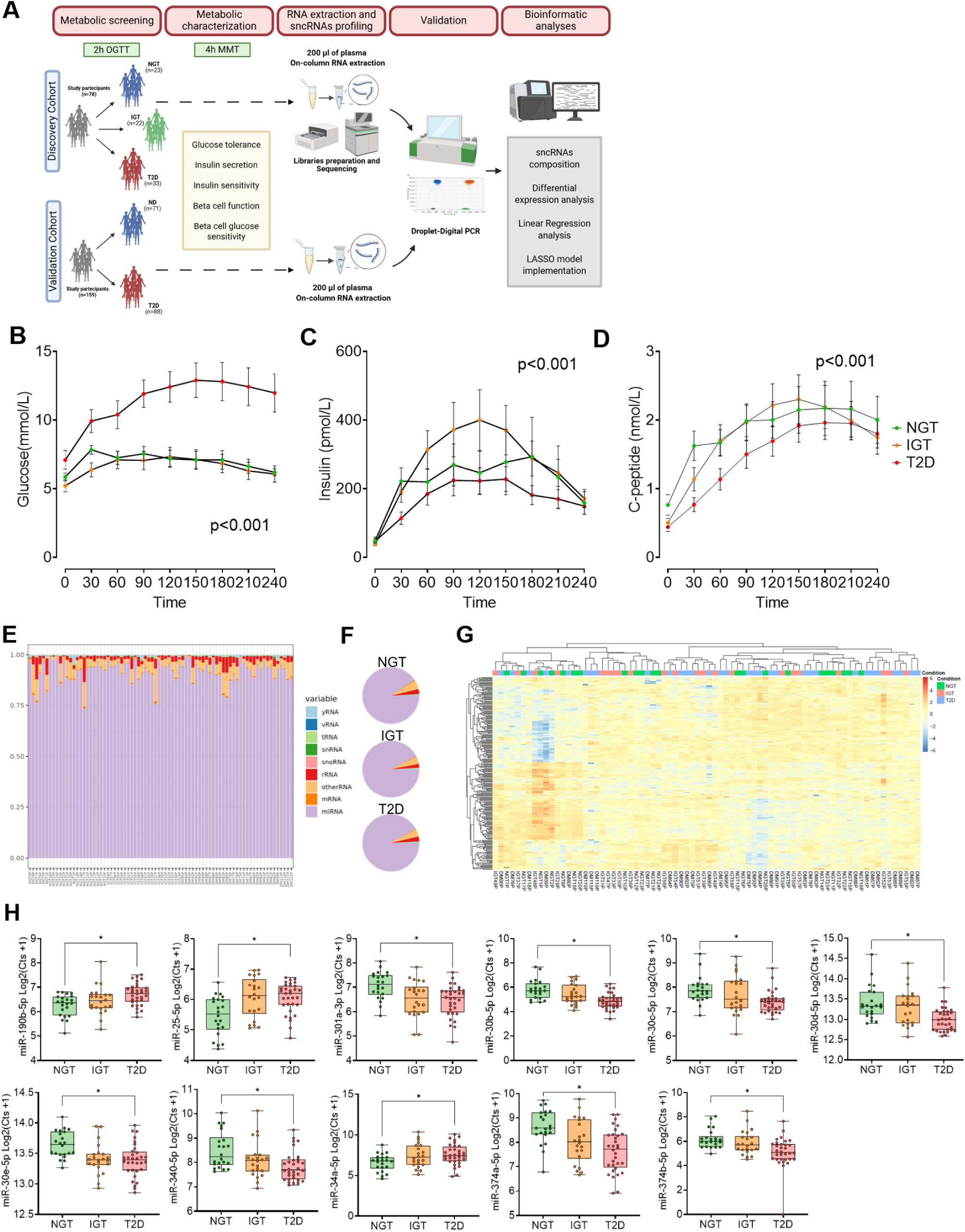
Circulating small non-coding RNA profiles and differential miRNA expression across normal glucose tolerance (NGT), impaired glucose tolerance (IGT) and type-2 diabetes (T2D) donors. **A.** Study design and experimental workflow. **B, C, D.** MMT curves of **(B)** Glucose, **(C)** Insulin, **(D)** C-peptide levels from NGT (green), IGT (orange) and T2D (red) individuals from the discovery cohort. *p≤0.05, was considered statistically significant for glucose, insulin and C-peptide levels for each time point. **E.** Stacked-bar plots of the relative abundance of sncRNA subtypes—yRNA, vRNA, tRNA, snRNA, snoRNA, mRNA, rRNA, miRNA and other RNA—normalized to total mapped reads in each individual sample (NGT, IGT, T2D). **F.** Pie charts showing the mean composition of sncRNA subtypes in plasma pooled by group, highlighting that miRNAs (lavender) comprise the majority of mapped sncRNA species in all metabolic states. **G.** Heatmap of hierarchical clustering (complete linkage, Euclidean distance) of 246 miRNAs with >5 count per million (CPM) in at least one sample; expression values were Z-score normalized across samples (blue = low, yellow = high). Top annotation bars denote metabolic status (green: NGT; orange: IGT; red: T2D), revealing distinct miRNA clusters associated with each group. **H.** Boxplots of log₂(CPM + 1) for eleven miRNAs significantly altered between NGT, IGT and T2D (miR-150-5p, miR-25-5p, miR-30a-3p, miR-30b-5p, miR-30c-5p, miR-93-5p, miR-122-5p, miR-223-3p, miR-374-5p, miR-423-5p). Boxes represent median and interquartile range; whiskers show minimum and maximum values; *p < 0.05 by one-way ANOVA with Tukey’s post hoc test.

The analysis of circulating small RNAs revealed, as expected, a predominant presence of miRNAs across all samples (**Figure 1E**), accounting for 90.8% in the NGT group, 92.5% in the IGT group, and 91.9% in the T2D group. The remaining small RNA classes collectively constituted less than 10% of the total mapped and assigned reads (**Figure 1F**). The average proportions of small non-coding RNA classes across the experimental groups are provided in **Supplementary Table 1**. Importantly, no significant differences were observed in the proportions of small RNAs among the three groups (**Figure 1F**).

Overall, we identified a total of 246 circulating miRNAs with more than 5 counts per million (CPM) in at least 50% of the samples within any group (NGT, IGT or T2D). To determine whether circulating miRNAs alone could distinguish between the three groups based on glucose tolerance stages, we conducted an unbiased unsupervised hierarchical clustering analysis of the miRNAs. The analysis revealed that circulating miRNAs, when considered collectively, do not effectively capture the subdivision of subjects based on their glucose tolerance stages (**Figure 1G**).

### A distinct set of circulating miRNAs is differentially expressed across individuals with NGT, IGT, and T2D

Since the overall miRNAs profile could not reliably distinguish between NGT, IGT, and T2D, we conducted a differential expression analysis to pinpoint specific miRNAs that might be associated with varying glucose tolerance conditions. This analysis revealed 11 miRNAs that were differentially expressed across the three groups (**Figure 1H**). Among these, 8 miRNAs were downregulated in the plasma samples of T2D patients compared to NGT subjects, including miR-301a-3p, several members of the miR-30 family (miR-30b-5p, miR-30c-5p, miR-30d-5p, and miR-30e-5p), miR-340-5p, miR-374a-5p, and miR-374b-5p. The remaining 3 miRNAs, miR-190b-5p, miR-25-5p, and miR-34a-5p, were upregulated in T2D samples compared to NGT (**Figure 1H**).

### Linear regression and LASSO models analyses revealed a three-miRNAs signature associated with beta cell dysfunction

Beta cell functional modelling encompasses two key measures: glucose sensitivity (GS) and rate sensitivity (RS). These parameters, derived from mathematical modelling of OGTT data, provide an estimate of complementary functional features of insulin secretion and are considered indexes of beta cell secretory function [2, 27]. Hence, these functional parameters can be utilized to estimate the association of circulating miRNAs to alterations of beta cell function. Therefore, we leveraged the comprehensive characterization of in vivo beta cell function performed in our cohort to identify relevant miRNAs associated with insulin secretion.

Firstly, we conducted a linear regression analysis between circulating miRNAs expression and the clinical/metabolic parameters (reported in **Table 1**) of the NGT, IGT and T2D individuals, after correction for age. Overall, we found 22 miRNAs significantly associated with RS and/or GS (**Figure 2A and Supplementary Table 2)**. Interestingly, multiple miRNAs showed significant association exclusively with RS and/or GS, while some of them showed significant associations with other parameters as well (i.e. Let-7c-5p, miR-1306, miR-30 family).

**Figure 2.**
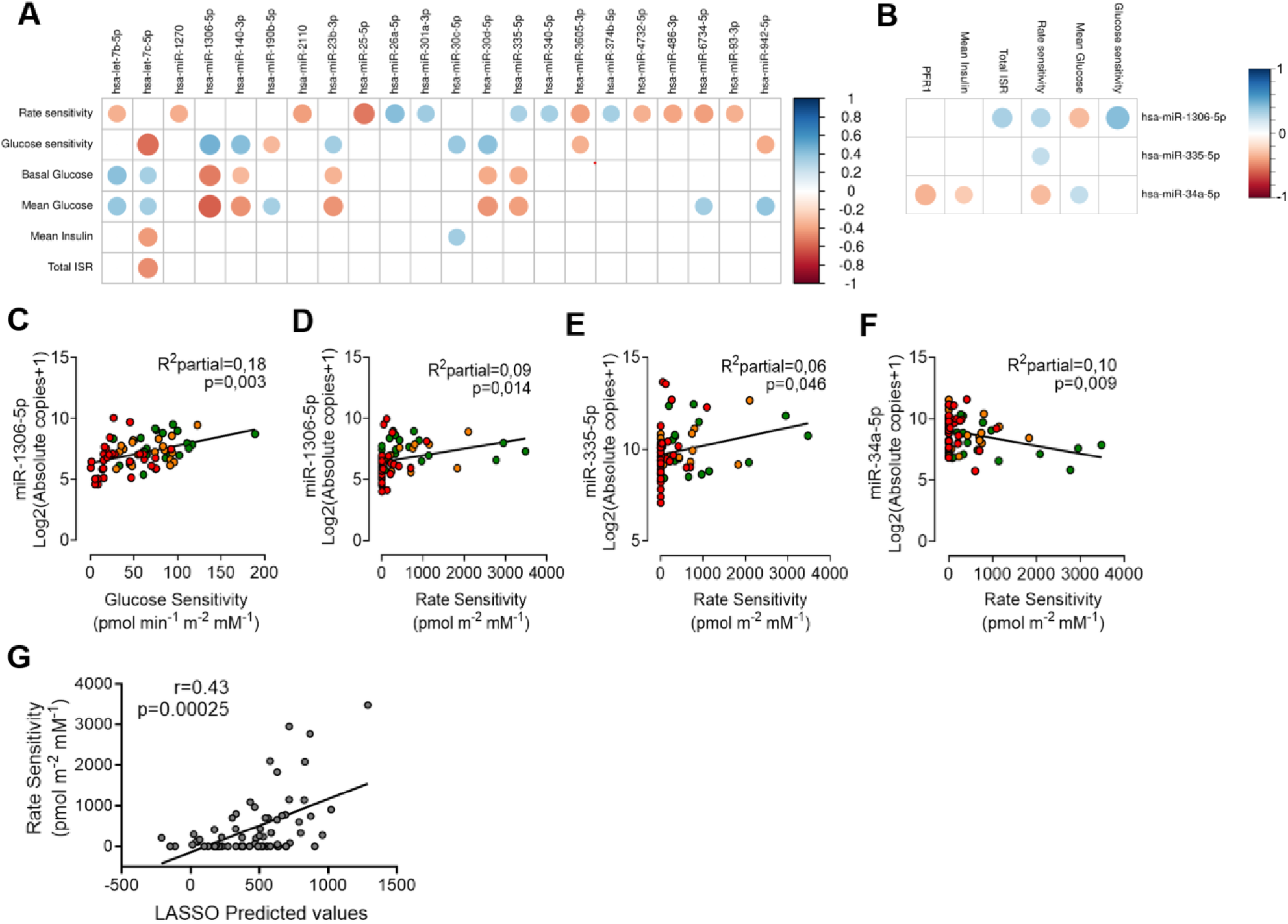
Circulating microRNAs reflect beta cell secretory dynamics and predict rate sensitivity via LASSO modelling. **A** Correlation plot of 22 plasma miRNAs (from small RNA-seq) versus OGTT-derived clinical and metabolic parameters - rate sensitivity (RS), glucose sensitivity (GS), potentiation factor ratio (PFR1), basal glucose, mean insulin concentration, total insulin secretion rate (ISR) and mean glucose-in 78 donors (23 NGT, 22 IGT, 33 T2D). Models were adjusted for age; only associations with p < 0.05 for the miRNA and beta cell functional measures (RS and/or GS) in multiple linear regression are shown. Dot area represents the significance of the association (wider= more significant); colour denotes coefficient sign (blue = positive; red = negative) of the √ (partial R²). **B.** Correlation plot of three ddPCR-validated miRNAs (hsa-miR-1306-5p, hsa-miR-335-5p and hsa-miR-34a-5p) versus a focused subset of OGTT parameters (PFR1, mean insulin, glucose sensitivity, rate sensitivity, mean glucose, total ISR), plotted as in (A). **C–F** Scatterplots of covariate-adjusted log₂ (ddPCR copies + 1) versus beta cell function parameters: **C.** miR-1306-5p vs. glucose sensitivity (pmol min^-1^ m^-2^ mM^-1^); **D.** miR-1306-5p vs. rate sensitivity (pmol m^-2^ mM^-1^). **E** miR-335-5p vs. rate sensitivity (pmol m^-2^ mM^-1^). **F.** miR-34a-5p vs. rate sensitivity. Data points represent individual donors (NGT: green; IGT: orange; T2D: red) with fitted linear regression. **G.** Observed versus LASSO-predicted rate sensitivity (pmol m^-2^ mM^-1^), data are shown as individual values. ISR, insulin secretion rate; NGT, normal glucose tolerance; IGT, impaired glucose tolerance; T2D, type 2 diabetes.

Secondly, to identify a robust and reliable circulating miRNAs signature linked to progressive beta cell dysfunction in T2D, we selected 19 candidates (**Supplementary Table 3**) based on at least one of these criteria: (*i*) a median UMI count > 100 (across all groups) coupled with a differential expression among glucose tolerance stages and/or (*ii*) median UMI count > 100 combined with significant associations (p < 0.05) to at least RS and/or GS beta cell-related metabolic parameters. These miRNAs were selected and further quantified by droplet digital PCR (ddPCR) to validate the results obtained using small RNA sequencing.

Notably, ddPCR analysis confirmed the progressive upregulation of miR-34a-5p across glucose tolerance stages (**Supplementary Figure 2A),** and demonstrated several correlations between miRNAs and beta cell functional parameters (**Figure 2B**). Interestingly, we found positive associations between miR-1306-5p and both GS (p = 0.003, partial R² = 0.18; **Figure 2C**) and RS (p = 0.014, partial R² = 0.09; **Figure 2D**). miR-1306-5p was also significantly associated with mean glucose and total insulin output (tISR) (**Supplementary Figure 2B, C**). Similarly, miR-335-5p was confirmed to be positively correlated with RS (p = 0.046, partial R² = 0.06; **Figure 2E**). Intriguingly, the analysis of ddPCR data showed a negative correlation between miR-34a-5p and RS (p = 0.009, partial R² = 0.10; **Figure 2F**), PFR1 (p = 0.003, partial R² = 0.12), and mean insulin levels (p = 0.04, partial R² = 0.06) (**Supplementary Figure 2D, E**), alongside with a positive correlation with mean glycemia (p = 0.038, partial R² = 0.06) (**Supplementary Figure 2F**). Collectively, these results indicate a three-miRNAs panel that is associated with key beta cell functional indexes.

Finally, given the pivotal role of RS as an early indicator of beta cell functional status, we applied a least absolute shrinkage and selection operator (LASSO) regression model to identify a composite biomarker signature predictive of RS. As predictors, we included ddPCR-quantified levels of miR-34a-5p, miR-335-5p, and miR-1306-5p, together with routinely available demographic and clinical variables (age, sex, BMI, fasting glucose, and 1-hour post-load glycemia from the OGTT). The final model retained the three miRNAs together with age and 1-hour post-load glycaemia from the OGTT as significant predictors of RS. When applied to our discovery cohort, predicted and observed RS values showed a significant positive correlation (Spearman’s ρ = 0.43; p = 2.5 × 10⁻⁴; **Figure 2G**), underscoring the model’s predictive accuracy. Together, these data suggest that a composite score incorporating miR-34a-5p, miR-335-5p, miR-1306-5p, age, and 1-hour glycemia can serve as a non-invasive early detector of beta cell secretion defects in individuals at risk for T2D.

### Validation of the three-miRNA signature in an independent cohort

To validate our three-miRNAs signature in an independent population, we recruited 158 subjects, 71 non-diabetic controls (39 females, 32 males; age 42 ± 13 years; BMI 26.2 ± 4.9 kg/m²) and 87 patients with established T2D (29 females, 58 males; age 60 ± 10 years; BMI 30.1 ± 5.1 kg/m²) (**Table 2**), who underwent MMT (**Figure 3A, B, C**) and calculation of functional parameters. Clinical and metabolic characteristics of this population are reported in **Table 2**. Glucose, insulin, and C-peptide levels during the MMT are shown in **Figure 3A, B, C**. According to GLMM with a Gamma distribution and log link, significant interactions between time and glucose tolerance were detected for all three markers (all p < 0.001), indicating distinct trajectories between groups over the course of the test. As expected, compared with non-diabetic individuals (ND), subjects with diabetes (T2D) exhibited significantly higher glucose concentrations (**Figure 3A**; p < 0.001), as well as higher insulin (**Figure 3B**; p < 0.001) and C-peptide (**Figure 3C**; p < 0.001) responses.

**Figure 3.**
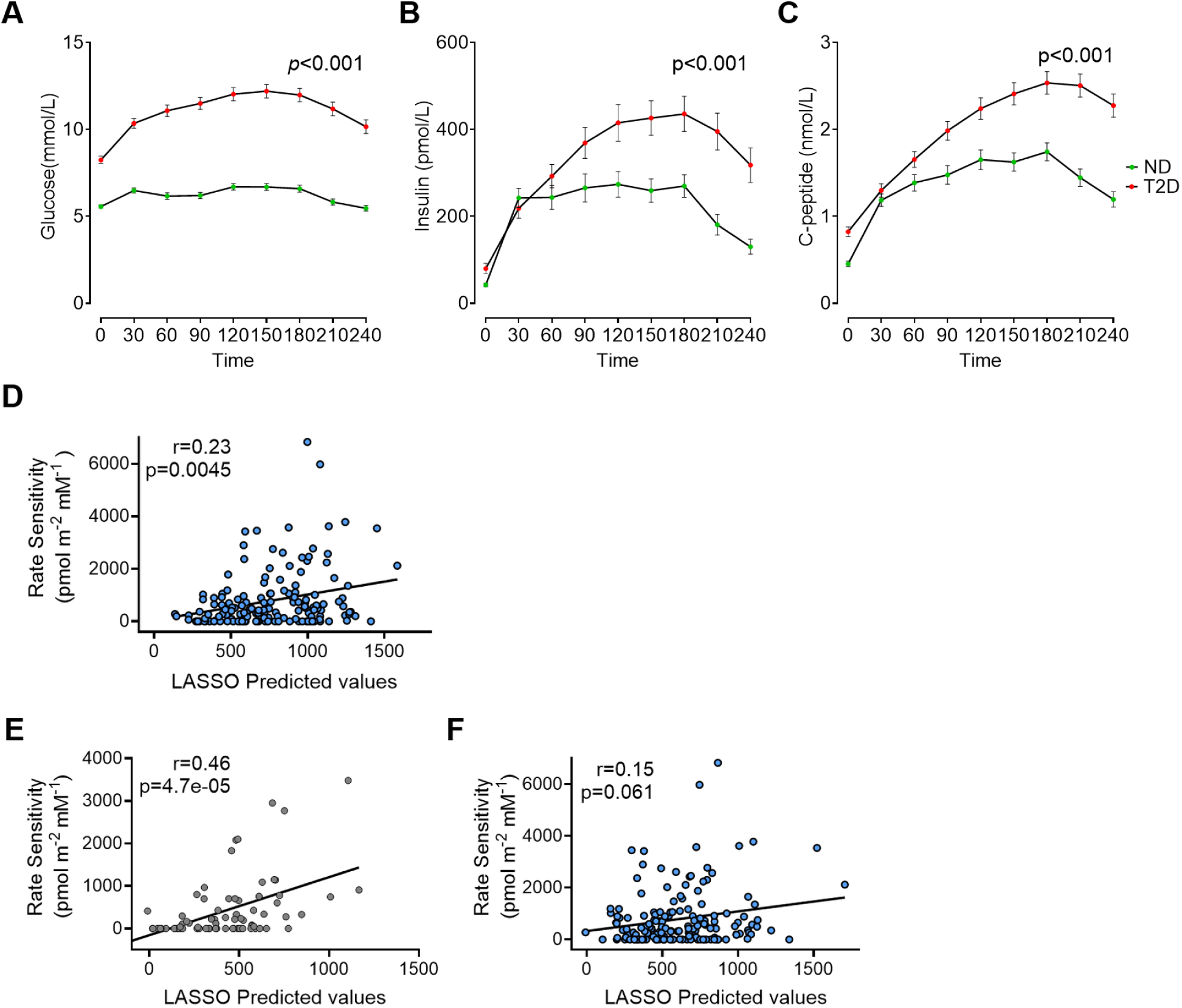
LASSO models integrating circulating miRNAs and glycaemic indices predict beta cell rate sensitivity. **A, B, C.** MMT curves from ND and T2D individuals from the discovery cohort. **(A)** Glucose, **(B)** insulin, **(C)** C-peptide levels in ND (green) and T2D (red). *p≤0.05, was considered statistically significant for glucose, insulin and C-peptide levels for each time point. **D.** Observed versus LASSO-predicted rate sensitivity (pmol m^-2^ mM^-1^) in the validation cohort (n=158 individuals) when including 60-min OGTT glycaemia (mmol/L) as the glycaemic covariate; Spearman r = 0.23, p = 0.0045. The solid line denotes the identity (y = x). **E.** Observed versus predicted rate sensitivity for the basal-glycaemia model in the training (discovery) cohort. **F.** Validation of the basal-glycaemia model in the validation cohort; data report observed versus predicted rate sensitivity. Data points represent individual participants; LASSO, least absolute shrinkage and selection operator; OGTT, oral glucose tolerance test; ddPCR, droplet digital PCR.

ND and T2D differed significantly in age (p<0.001) and BMI (p<0.001), as well as in all metabolic parameters. Beta cell function parameters also differed significantly, with the T2D group displaying higher bISR (104.94 ± 63.59 pmol min⁻¹ m⁻² vs 60.53 ± 32.30 pmol min⁻¹ m⁻², p<0.001) and tISR (69.22 ± 32.68 nmol m^-2^ vs 47.92 ± 22.50 nmol m^-2^, p<0.001), but lower RS (454.05 ± 628.71 pmol m^-2^ mM^-1^ vs 1207.83 ± 1391.13 pmol m^-2^ mM^-1^ p<0.001), GS (61.06 ± 65.86 pmol min^-1^ m^-2^ mM^-1^ vs 97.13 ± 66.70 pmol min^-1^ m^-2^ mM^-1^, p<0.001) and PFR1 (1.27 ± 0.61 vs 1.44 ± 0.70, p=0.02) values.

Then, we measured miR-34a-5p, miR-335-5p, and miR-1306-5p in the plasma of all subjects by ddPCR. After adjusting for age, sex, and BMI, miR-34a-5p remained significantly elevated in T2D (**Supplementary Figure 3**), underscoring its robustness as a marker of deteriorating glucose tolerance.

We next assessed whether the LASSO model derived in the discovery cohort could be used to predict RS in an independent validation set. For each individual in the validation cohort, predicted RS values were obtained by applying the fixed coefficients of the discovery LASSO model to the log2-transformed ddPCR expression levels of the three selected miRNAs (miR-34a-5p, miR-335-5p, and miR-1306-5p), together with age and 1-hour post-load glycemia as input variables. Predicted RS values showed a significant positive association with observed RS measurements (Spearman’s ρ = 0.23, p = 0.0045; **Figure 3D**), supporting the ability of this composite signature to identify early beta cell secretory dysfunction in an independent population.

To enhance the clinical applicability of the model, we fitted a simplified LASSO model in the discovery cohort, replacing 1-hour post-load glycemia with fasting plasma glucose while retaining age and the three miRNA expression levels as candidate predictors. The penalty parameter (λ) was selected by cross-validation within the discovery cohort, and the resulting coefficients were fixed. In this simplified model, predicted RS values showed a moderate positive correlation with observed RS in the discovery cohort (Spearman’s ρ = 0.46, p = 4.7 × 10⁻⁵; **Figure 3E**). When the locked simplified model was applied to the independent validation cohort (n = 158), predicted RS values remained positively associated with observed RS (Spearman’s ρ = 0.15, p = 0.06; **Figure 3F**), indicating a consistent, albeit weaker, predictive signal and supporting the potential utility of this composite marker. Importantly, replacing 1-hour post-load glycemia with fasting glucose enables the use of this predictive model without the need for an OGTT. This adjustment increases clinical convenience, reduces patient burden, and facilitates broader application of the miRNA-based signature in routine care settings.

## Discussion

Our study demonstrates that circulating miRNAs can accurately mirror early functional defects of pancreatic beta cells in humans. By integrating plasma miRNA profiling with model-based measures of insulin secretion, we identified a robust three-miRNAs signature (miR-34a-5p, miR-335-5p, and miR-1306-5p) that reflects in vivo beta cell dysfunction in individuals across the spectrum of glucose tolerance. This finding provides a biological and clinically accessible link between systemic miRNA levels and the dynamic capacity of beta cells to respond to metabolic demands, offering new insights into the molecular determinants of early diabetes progression.

The decline in beta cell function is a key event in the natural history of T2D [28], yet its direct assessment requires complex physiological tests such as the OGTT or the MMT, which are difficult to implement in large-scale or longitudinal studies. Hence, easily accessible and measurable biomarkers reflecting beta cell dysfunction are urgently needed to improve early diagnosis and improve therapeutic outcomes.

Our results bridge this gap by showing that a small set of circulating miRNAs, identified using small RNA-seq and digital droplet PCR validation from two independent cohorts of living donors subjected to detailed metabolic profiling, can capture critical aspects of beta cell secretory dynamics derived from mathematical modelling, including RS, a parameter reflecting the promptness of insulin release to rising glucose. MiRNAs are critical regulators of various beta cell functional processes [29] and several blood circulating miRNAs have been consistently linked to pre-diabetes, T2D and its complications [20]. Hence, certain miRNAs consistently emerged as potential biomarkers for the diagnosis, prognosis and progression of diabetes [30][31], and some of them were also linked to therapeutic outcomes [32].

In this context, our study encompasses a unique workflow to identify novel circulating biomarkers of early and established beta cell dysfunction in the progression of T2D. First, all the blood samples subjected to small RNAs profiling were processed according to a stringent SOP [25] to reduce the overall small RNAs/miRNAs variance introduced by multiple preanalytical biases.

Second, the study design included two independent cohorts composed of 78 individuals (discovery cohort: 23 NGT, 22 IGT, 33 T2D) and 158 individuals (validation cohort: 71 ND, 87 T2D); notably, all the individuals underwent a deep metabolic profiling which included an accurate mathematical modelling of OGTT or MMT data to retrieve unique indexes of beta cell function, including GS and RS. RS quantifies early-phase insulin release and provides a direct measure of beta cell function that mirrors the characteristic reduction in first-phase insulin secretion typically seen in T2D [33, 34]. Further, we previously demonstrated that reduced GS and RS can together predict diabetes appearance in a cohort of non-diabetic patients after ∼50% acute removal of total pancreatic beta cell mass [4]. Therefore, the routine adoption of RS would be ideal to track the progression of T2D. However, RS use in the clinical practice is currently limited, as its measurement requires complex mathematically modelling of the time-sequentially measured C-peptide and glucose concentrations under various conditions that stimulate beta cells, such as MMT and OGTT [35]. Consequently, using RS as a benchmark for the discovery of novel biomarkers may offer particular advantages to identify novel surrogate markers of beta cell function.

Third, we applied an unbiased machine-learning approach (LASSO) to select miRNAs alongside simple and accessible demographic and clinical variables, thereby identifying both molecular and clinical features for a composite biomarker signature of beta cell function.

The final model incorporated expression levels of a three-miRNAs signature (miR-34a-5p, miR-1306-5p, and miR-335), together with age and 1-hour post-OGTT glycemia, to predict RS. LASSO prioritized 1-hour glycemia rather than fasting glucose; this was expected, since 1-hour plasma glucose serves as a practical clinical index of metabolic impairment and better identifies individuals with severe insulin resistance and impaired beta cell function [36][37]. The predicted RS values from the statistical model showed a significant positive correlation with the observed values (ρ = 0.43, p = 2.5×10^-4^), indicating moderate predictive accuracy. Importantly, the same model [miR-34a-5p + miR-1306-5p +and miR-335 + Age (y) + 1-hour glucose levels (mM)] was also applied to the validation cohort (n=158 individuals), confirming a significant positive correlation between predicted and observed RS values (p = 0.0045; ρ = 0.23), indicating good prognostic capability of beta cell function though RS prediction.

In the final model, the three miRNAs were selected using two distinct methodologies across sequential analysis of the discovery and validation cohorts, comprising a total of 236 individuals fully metabolically characterised. Hence, in this context, the model was subjected to a wide validation across two independent cohorts. Nevertheless, the modest, though significant, correlation coefficients observed both between individual miRNA levels and beta cell functional metrics and between LASSO-predicted values and RS, indicate that additional factors likely contribute to the remaining unexplained variance. Importantly, even in its current form, the inclusion of miRNAs offers added value as they serve as informative, complementary biomarkers alongside established clinical indicators. Moreover, substituting 1-hour OGTT glycaemia with fasting plasma glucose in our model still produced a robust, statistically significant prediction of RS in the discovery cohort (ρ = 0.46, p = 4.7 × 10⁻⁵) and a similar association in the validation cohort (ρ = 0.15, p = 0.06). The attenuation of effect size in the validation cohort suggests that the simplified model captures only part of the variability in RS and warrants further refinement and validation in larger cohorts. However, by relying on a single fasting measurement rather than a full OGTT, this modification greatly improves clinical feasibility while preserving predictive performance. We therefore encourage testing this streamlined model in additional independent cohorts to pave the way for its adoption in clinical routine testing. Among the three identified miRNAs, miR-34a-5p emerged as a central node linking metabolic stress to beta cell dysfunction.

MicroRNA miR-34a-5p was consistently upregulated in the plasma of individuals with T2D across both discovery and validation cohorts, thus in line with multiple previous reports [20]. miR-34a-5p is broadly expressed in multiple cells and tissues and shows dysregulation in T2D [38–41]. Intriguingly, in pancreatic beta cells, inflammatory and lipotoxic stimuli drive miR-34a upregulation [42, 43] which in turn impairs insulin granule exocytosis, reduces glucose-stimulated insulin secretion, and promotes apoptosis [42, 43]. Our findings of elevated plasma levels of miR-34a-5p, and its inverse correlation with rate sensitivity, PFR1, and mean insulin levels, aligns with these mechanistic insights, thus potentially suggesting a putative beta cell origin.

Similarly, miR-335 was also previously reported to be associated with T2D in multiple contexts [44–47]. Notably, in both the human beta cell line EndoC-βH1 and rat INS-1 cells, miR-335 overexpression markedly reduced glucose-stimulated insulin secretion. Mechanistically, this effect originates from miR-335-mediated downregulation of key exocytotic proteins SNAP25, Syntaxin-binding protein 1 (STXBP1), and Synaptotagmin-11 (SYT11), which impairs the priming of insulin granules at the plasma membrane and reduce first-phase insulin release [48]. Given its direct impact on early insulin secretion dynamics, the inclusion of miR-335 in our composite signature for predicting RS is both mechanistically and clinically justified. The inclusion of miR-1306-5p, less characterized in the endocrine pancreas but showing a consistent association with glucose sensitivity, suggests that under conditions of increased insulin demand, this miRNA may participate in the modulation of the early phase of insulin secretion. Together, these three molecules capture distinct yet converging facets of beta cell failure: survival stress, exocytotic machinery impairment, and defective glucose sensing.

In conclusion, we provide direct evidence that a three-miRNAs signature (miR-34a-5p, miR-1306-5p, and miR-335) can serve as a sensitive biomarker of early beta cell dysfunction. While further validation in larger cohorts is necessary to confirm the clinical reliability of this predictive signature, our results support a model in which metabolic stress triggers coordinated miRNA responses that mirror the progressive decline of beta cell competence, underscoring the feasibility of using circulating miRNAs, in combination with easily obtainable clinical parameters, to assess beta cell function and identify individuals at risk of developing diabetes. Future longitudinal and interventional studies will be essential to determine whether modulation of these miRNAs reflects or even influences the reversibility of beta cell failure in T2D, enhancing the possibility of monitoring of beta cell dysfunction in clinical settings.

## Supporting information

ESM

## Data Availability

Raw and analysed data are available from the corresponding author upon request.

## ACKNOWLEDGEMENTS

We acknowledge the help of Stefano Auddino for the analysis of small RNA seq data.

## CONTRIBUTION STATEMENT

EA and GS contributed to the conceptualisation, supervision, and coordination of the study, the design of the methodology, conducted the investigation and implemented the bioinformatics and statistical analysis, composed the figures and write and review the manuscript.

GEG, MB, GG, DF, GDG, GC, AD and MP conducted the experiments and the analyses and interpreted the data. GQ and SA contributed to the investigation and to the design of the methodology. AG, TM and GD contributed to the investigation, to the design of the methodology and helped acquire funding for the research. AM and RR contributed to the investigation and to the design of the methodology. FD and GS contributed to the conceptualisation, supervision and coordination of the study and the design of the methodology, write and review the manuscript and helped acquire funding for the research.

FD, GS, TM and GD are the guarantors of this work and, as such, had full access to all the data in the study and takes responsibility for the integrity of the data and the accuracy of their analysis.

All authors substantially contribute to the conception or design of the work, or acquisition, analysis or interpretation of data of the work. All authors approved the final version.

## ABBREVIATIONS

GS: Glucose Sensitivity
IGT: Impaired Glucose Tolerance/Tolerant
MiRNAS: MicroRNAs
MMT: Mixed Meal Test
ND: Non-Diabetic
NGT: Normal Glucose Tolerance/Tolerant
PFR1: Potentiation Factor Ratio
RS: Rate Sensitivity
T2D: Type 2 Diabetes
OGTT: Oral Glucose Tolerance Test

