## Supplementary material for "Plasma Small RNA profiling reveals a predictive three-miRNAs signature for early beta cell dysfunction across glucose tolerance stages": ESM

**Electronic Supplementary Material (ESM)**

#### ESM Methods

##### Study population and metabolic profile

Seventy-eight subjects (40 males; 38 females; mean age 66.6±10.7 [years ± SD]) undergoing pylorus-preserving pancreatoduodenectomy were recruited from January 2017 to July 2019 at the Digestive Surgery Unit and studied at the Centre for Endocrine and Metabolic Diseases unit (Agostino Gemelli University Hospital, Rome, Italy) and underwent a complete metabolic screening including Oral Glucose Tolerance Test (OGTT) or a Mixed Meal Test (MMT). Based on OGTT in the days immediately before surgery, subjects were classified as normal glucose tolerant (NGT= 23), impaired glucose tolerant (IGT=22) and type 2 diabetic (T2D) with disease onset longer than 2 years (n=33). Among diabetic patients, we selected those with newly stable metabolic control (HbA1c <7.0 %) on diet alone or diet + metformin. All 78 subjects underwent both an OGTT and/or MMT to evaluate insulin secretion (from C-peptide deconvolution) (**Table 1**).

In addition, to validate our findings, we enrolled 158 subjects without pancreatic diseases, who attended the Centre for Endocrine and Metabolic Diseases unit (Agostino Gemelli University Hospital, Rome, Italy) and the Diabetes outpatients clinic (Azienda Ospedaliero-Universitaria Pisana, Pisa, Italy) between July 2022 and May 2024. The subjects were divided into 71 non-diabetic controls, ND, (39 women, 32 men) and 87 patients with established T2D (29 women, 58 men), who underwent MMT and calculation of functional parameters (**Table 2**). Among diabetic patients, we selected those with stable metabolic control or with secondary failure metformin monotherapy (HbA1c>7.0% and <9%) in metformin monotherapy.

The study protocol (ClinicalTrials.gov NCT02175459) was approved by the Ethical Committee Fondazione Policlinico Universitario Agostino Gemelli IRCCS – Università Cattolica del Sacro Cuore (P/656/CE2010 and 22573/14), and all participants provided written informed consent, followed by a comprehensive medical evaluation.

**Oral Glucose Tolerance Test and Mixed Meal Tests (MMT)**

A standard 75 g oral glucose tolerance test was performed with measurement of glucose, insulin and C-peptide at 0, 30, 60, 90, 120 min after glucose load. Based on the pre-surgery OGTT results, we classified the patients according to the ADA classification (American Diabetes Association, 2019). subjects whose 2 h post glucose load was below 140 mg/dl were defined as normal glucose tolerant (NGT), subjects whose 2 h post glucose load was 140–199 mg/dl were defined as impaired glucose tolerant (IGT) and subjects whose 2 h post glucose load was higher than 200 mg/dl and with known history of type 2 diabetes of over two years and/or on anti-diabetic medications, were defined as diabetic (T2D).

A mixed meal test (MMT) was performed, as previously described [1]. Patients were instructed to consume a meal of 830 kcal (107 kcal from protein, 353 kcal from fat, and 360 kcal from carbohydrates) within 15 min. Blood samples were drawn twice in the fasting state and at 30 min intervals over the following 240 min (sample time 0', 30', 60', 90', 120', 150', 180', 210' and 240') for the measurement of plasma glucose, insulin, C-peptide. Insulin levels were determined using a commercial RIA kit (Medical System, Immulite DPC, Los Angeles, CA). Plasma glucose concentrations were determined by the glucose oxidase technique, using a glucose analyzer (Beckman Instruments, Palo Alto, CA, USA). Plasma C-peptide was measured by auto DELPHIA automatic fluoroimmunoassay (Wallac, Turku, Finland), with a detection limit of 17 pmol/L.

During the OGTT and MMT, insulin secretion was derived from C-peptide levels by deconvolution and functional parameters [Beta-cell Glucose Sensitivity (GS), Rate sensitivity (RS), basal and total Insulin Secretion Rate (bISR and tISR) and potentiation factor ratio (PFR1)] were estimated by mathematical modelling, as previously described [2] [2–4]. The assessed parameters included: GS, i.e. the slope of the static relationship between insulin secretion and glucose concentration; RS which reflects early phase insulin release; bISR and tISR representing the basal insulin secretion and the total insulin output, respectively ; potentiation factor ratio (PFR1), reflecting the enhancement of insulin secretion over time, due to multiple mechanisms such as sustained hyperglycaemia, non-glucose substrates, incretin effect and neural influences [2], [3], [4]. For the discovery cohort, calculation of parameters was obtained from OGTT measurements; calculation of RS, the parameter to be predicted by LASSO model analysis (indicated in **Statistical Analysis** section), the corresponding values derived from MMT were employed for those subjects in which OGTT data were missing.

**Blood samples collection, plasma samples processing and total RNA extraction**

Blood samples were collected following a specific Standard operating Procedure (SOP) as previously reported [5], [6]. Blood samples were collected in 3.5 mL $K_2$-EDTA tubes, inverted ten times, and stored upright at room temperature (15-25°C) before being processed within 2 hours from collection. The initial centrifugation was carried out at 1800xg for 10 minutes at 15-25°C to separate the blood cells from the plasma. The plasma was then carefully transferred into smaller, sterile, apyrogenic, and nuclease-free tubes (2 mL) without disturbing the white blood cell interface, leaving a 2-3 mm layer of plasma above the leukocytes. A second centrifugation was performed at 1200xg for 20 minutes at 10°C to remove any remaining contaminant cells and platelets. Subsequently, multiple 200 µL plasma aliquots (up to 5 aliquots per sample, where possible) were stored in nuclease-free tubes at -80°C before being transferred to the analytical laboratory for further analysis.

Total RNA extraction was performed from 200 µL of plasma through Serum/Plasma Norgen kit (cat. 55000, Thorold, ON L2V 4Y6, Canada) following manufacturer's recommendation

**Small RNA cDNA library preparation and QC**

Small RNA-derived cDNA libraries were prepared using QiaSeq miRNA library kit (cat. 331505, Qiagen) as previously described [5], [6]. QIASeq strategy assign Unique Molecular Index (bound to reverse transcription primers) during reverse transcription step to every mature miRNA molecule, to enable unbiased and accurate miRNome-wide quantification of mature miRNAs by NGS. A total of

5 uL of total RNA extracted from 200 uL of plasma (see above) was used as an input for QiaSeq miRNA library kit and then processed following manufacturers' recommendation and as reported in Grieco et al. 2021 [5]. Then, libraries quality control (QC) was performed quantifying their concentration through QUBIT 3.0 spectrofluorometer (Qubit™ dsDNA HS Assay Kit, cat. Q32854, Thermofisher Scientific) and assessing their quality using capillary electrophoresis in Bioanalyzer 2100 (Agilent High Sensitivity DNA kit cat. 5067-4626, Thermofisher Scientific). High quality of libraries was evaluated considering electropherograms showing a peak comprised between 175 and 185 bp. Following QC, all libraries were normalized until 2 nM and pooled, denatured in 0.2 N NaOH and further sequenced (final concentration 175 pM) on Illumina NovaSeq 6000 platform [NovaSeq 6000 SP Reagent Kit (100 cycles) cat. 20027464, NovaSeq XP 2-Lane Kit cat. 20021664, Illumina] using the XP protocol applying 75x1 single reads. Data were returned from BaseSpace Sequence Hub as demultiplexed FASTQ files.

**Small RNAs Quantification and Profiling**

FastQ files obtained from the small RNA sequencing were analyzed with the sRNAbench online pipeline. Reads were processed with the Qiagen (with UMIs) protocol for adapters and duplicates removal. Reads were mapped in genome mapping mode, using the Human reference Genome Reference Consortium Human Build 38 patch release 13 (GRCh38.p13). Alignment was performed with the bowtie algorithm using the seed option with length (L) = 20, maximum number of mismatches (N) = 1, minimum length of the read =15, minimum number of read count =2 and maximum number of multiple mappings equal to 10. Mapped reads were annotated to miRNAs using miRBase release 22.1. The raw miRNA counts matrix was obtained from the *mature_sense.grouped* file of each sample generated from sRNAbench. Mapped reads were also annotated to the other sRNAs using different annotation databases (RNAcentral database 20.0, ncRNA from Ensembl release 104 and cDNA from Ensembl release 104) and classified as yRNA, vRNA, tRNA fragments, snRNA, snoRNA, rRNA, mRNA fragments and other RNA. After the quantification step, sRNA profiling was performed using the *mappingStat.txt* file from each sample, to compute the number of reads annotated to the different sRNAs classes.

**Selection of miRNAs of interest for the validation stage**

To select the most relevant circulating microRNAs to be included in the validation stage, we adopted the following criteria: (i) differentially expressed miRNAs across glucose tolerance groups with a median UMI (Unique Molecular Identifier) count from sequencing of 100 or more, and/or (ii) miRNAs with a median UMI count of 100 or more with almost one significant association with GS and/or RS as parameters indexes of beta cell function.

**Reverse Transcription and ddPCR**

Validation of selected miRNAs identified, through both differential expression analysis and regression analysis, was performed through miRCURY LNA Reverse Transcription and subsequent droplet digital PCR (ddPCR) detection.

In details, their expression was analysed in all plasma samples using miRCURY LNA assay primers (Qiagen) through a standardised protocol. RNA (the same used for small RNA sequencing) was reverse transcribed. Briefly, 6 µL of RNA were added to 4 µL of 5X miRcury SYBR Green RT Reaction Buffer, 2 µL of 10X miRcury RT enzyme mix and 8 µL $H_2O$. The reaction product was incubated at 42 °C for 60 min and then 95 °C for 5 min. Then, droplet digital PCR was performed on a BioRad QX200 system using a EvaGreen assay (BioRad, Mississauga, ON, Canada). Each PCR reaction contained 11 µL of QX200 Evagreen Supermix, 1.1 µL of each 20X Qiagen miRCURY LNA PCR assay, 5.9 µL of H2O and 4 µL of template cDNA in a final volume of 22 µL. The PCR reactions were mixed, centrifuged briefly and 20 µL transferred into the sample well of a DG8™ cartridge. After adding 70 µL of QX200™ droplet generation oil into the oil wells, the cartridge was covered using a DG8™ gasket, and droplets generated using the QX200™ droplet generator. Droplets were carefully transferred into PCR plates using a multi-channel pipette and the plate sealed using PCR plate heat seal foil and the PX1™ PCR plate sealer. PCR was performed in a SimpliAmp touch thermal cycler (Thermofisher Scientific). The PCR protocol was 95°C for 10 min; 45 cycles of: 95°C for 30 s, optimal annealing temperature (54 °C for miR-34a-5p and miR-335-5p or 56°C for miR-1306-5p); 4°C for 5 min and 90°C for 5 min. PCR plates were transferred into a QX200™ droplet reader to count positive and negative droplets. Thresholds to separate positive from negative droplets were set manually for each miRNA using the histogram function and reads analysed using QuantaSoft™ Analysis Pro software (Version 1.2, BioRad, Mississauga, ON, Canada).

**Statistical Analysis**

In both cohorts (discovery and validation), baseline clinical characteristics were compared across glucose tolerance groups (NGT, IGT, and T2D). For each variable, a linear model was initially fitted, and the normality of residuals was assessed using the Shapiro–Wilk test. When residuals were normally distributed, overall group differences were evaluated using one-way ANOVA, followed by Tukey's test for pairwise comparisons. For variables with non-normally distributed residuals, the non-parametric Kruskal–Wallis test was applied, and Dunn's test was used for post hoc comparisons.

To analyze longitudinal data from Mixed Meal Tests (MMT) or Oral Glucose Tolerance Test (OGTT), Generalized Linear Mixed Models (GLMMs) were implemented for glucose, insulin, and C-peptide concentrations. Each marker was modeled as the dependent variable, with time and glucose tolerance status (Normal Glucose Tolerance, Impaired Glucose Tolerance, Type 2 Diabetes) as fixed effects, and subject ID as a random intercept to account for repeated measurements. Given the skewness and strictly positive nature of the data, a Gamma distribution with a log link was used. Type III Wald chi-square tests were performed to assess the main effects of time, glucose tolerance, and their interaction. Observed mean values with standard error bars were plotted over time to visualize glucose, insulin, and C-peptide concentrations during the MMT for each group.

Regarding the Small RNA sequencing data analysis, after the quantification step, the total amount of reads assigned to miRNAs for each sample was computed. Samples with <1.000.000 miRNA reads were removed from the further analyses (n=2 samples removed). After the removal of samples with low miRNAs expression, the next step was the low counts filtering to remove miRNAs with low expression. MiRNA read counts were converted in Counts per Million (CPM), and features with >5 CPM in at least 50% of samples were maintained, resulting in the identification of n=246 miRNAs. Filtered miRNA counts were normalized with DESeq2 median of the ratio's method, to account for differences in RNA composition and library depth. Pairwise Differential Expression analysis between the different group of patients identified according to glucose tolerance was performed with DESeq2's Wald test. The analysis was performed with the age as covariate. P-values were corrected for multiple test with Benjamin Hochberg procedure (padj). MiRNAs with padj<0.05 were considered as differentially expressed between the two groups tested.

Validation of miRNAs 'differential expression detected using sequencing was further evaluated with droplet digital PCR (ddPCR) data using linear models. Copies were log2 scaled after the addition of a pseudo count. A linear model with the log2 scaled copies as dependent variable and the stratification and age as independent variables were fitted for each miRNA. Pairwise comparison between the different groups were performed. P-values <0.05 associated to the group identify a miRNA differentially expressed between the two Stratifications, after correcting for Age.

Associations between miRNA expression levels (from both sequencing and ddPCR data) and clinical parameters were assessed using linear regression models. Raw expression values were log2-transformed after adding a pseudo-count to avoid issues with zero values. To reduce the influence of potential outliers on model estimates, Cook's distance was calculated for each observation in each model. Observations with Cook's distance greater than five times the average Cook's distance within the model were removed before model fitting.

Linear models were then re-estimated for each miRNA–clinical parameter pair. Associations were considered statistically significant when the p-value of the coefficient assigned to the clinical parameter was $< 0.05$, irrespective of covariate (age). The partial $R^2$ of each clinical parameter was estimated using the partial_r2() function from the sensemakr R package. To retain directional information, the signed partial R was calculated as the square root of the partial $R^2$, multiplied by the sign of the coefficient. For visualization purposes, log2-transformed expression values were adjusted for the effects of covariates. This adjustment was performed by estimating the predicted contribution of each covariate (coefficient × covariate value) for each data point, summing these contributions, and subtracting the total from the normalized log2 expression values. This yielded covariate-adjusted expression levels reflecting the effect of the clinical parameter of interest.

To identify a putative miRNA-based signature predictive of beta cell function, a LASSO regression model was implemented using rate sensitivity as the clinical outcome. The log2-transformed copy numbers of miR-34a-5p, miR-1306-5p, and miR-335-5p, along with age, sex, BMI, 1-hour glycemia, and basal glycemia, were included as candidate predictor variables. Model tuning and selection of the optimal regularization parameter (lambda) were performed using leave-one-out cross-validation (LOOCV) with the cv.glmnet() function from the glmnet package. The value of lambda that minimized the mean squared error (MSE) was used to fit the final model. The selected model retained all three miRNAs, age, and 1-hour glycemia as predictors of rate sensitivity. Predictive performance was evaluated by calculating the correlation between predicted and observed values of rate sensitivity in the discovery cohort, yielding a statistically significant positive association (Spearman's $\rho=0.43$, $p = 2.5 \times 10^{-4}$), indicating that the model reliably captures variability in beta cell function.

### ESM Tables

**Supplementary Table 1.** Classes of small RNAs (represented as percentage of the total with standard deviation) present in plasma samples from NGT, IGT, and T2D subjects.

| sncRNA class | NGT | IGT | T2D |
|---|---|---|---|
| *mRNA* | 0.406 ± 0.285 | 0.321 ± 0.253 | 0.333 ± 0.206 |
| *miRNA* | 90.75 ± 6.969 | 92.531 ± 4.966 | 91.915 ± 4.713 |
| *otherRNA* | 5.097 ± 3.692 | 4.128 ± 3.256 | 4.205 ± 2.617 |
| *rRNA* | 2.796 ± 3.226 | 2.093 ± 1.374 | 2.689 ± 1.848 |
| *snRNA* | 0.042 ± 0.033 | 0.11 ± 0.139 | 0.086 ± 0.135 |
| *snoRNA* | 0.027 ± 0.016 | 0.033 ± 0.017 | 0.039 ± 0.027 |
| *tRNA* | 0.26 ± 0.186 | 0.319 ± 0.252 | 0.315 ± 0.198 |
| *vRNA* | 0.002 ± 0.002 | 0.003 ± 0.004 | 0.002 ± 0.001 |
| *yRNA* | 0.619 ± 0.883 | 0.461 ± 0.436 | 0.416 ± 0.359 |

**Supplementary Table 2**. Table reporting linear regression analysis results (p value and partial R) of 22 miRNAs identified by sequencing significantly associated with at least one clinical parameter among those directly offering a relevant measure of beta cell function (RS and/or GS).

| miRNA | Clinical parameter | p | Partial R |
|---|---|---|---|
| *hsa-let-7b-5p* | Rate sensitivity | 4,65E-03 | -3,55E-01 |
| | Basal Glucose | 8,58E-04 | 4,13E-01 |
| | Mean Glucose | 2,15E-03 | 3,83E-01 |
| *hsa-let-7c-5p* | Glucose sensitivity | 2,64E-06 | -5,57E-01 |
| | Mean Insulin | 5,06E-04 | -4,26E-01 |
| | Basal Glucose | 7,19E-03 | 3,38E-01 |
| | Mean Glucose | 5,41E-03 | 3,44E-01 |
| | Total ISR | 1,03E-04 | -4,70E-01 |
| *hsa-miR-1270* | Rate sensitivity | 3,40E-03 | -3,66E-01 |
| *hsa-miR-1306-5p* | Mean Glucose | 3,51E-07 | -5,90E-01 |
| | Basal Glucose | 1,70E-05 | -5,17E-01 |
| | Glucose sensitivity | 1,01E-04 | 4,74E-01 |
| *hsa-miR-140-3p* | Glucose sensitivity | 6,24E-04 | 4,23E-01 |
| | Mean Glucose | 2,27E-04 | -4,52E-01 |
| | Basal Glucose | 9,68E-03 | -3,29E-01 |
| *hsa-miR-190b-5p* | Mean Glucose | 7,02E-03 | 3,36E-01 |
| | Glucose sensitivity | 9,95E-03 | -3,22E-01 |
| *hsa-miR-2110* | Rate sensitivity | 8,38E-04 | -4,10E-01 |
| *hsa-miR-23b-3p* | Glucose sensitivity | 6,27E-03 | 3,43E-01 |
| | Basal Glucose | 5,70E-03 | -3,44E-01 |
| | Mean Glucose | 2,70E-04 | -4,44E-01 |
| *hsa-miR-25-5p* | Rate sensitivity | 9,62E-06 | -5,22E-01 |
| *hsa-miR-26a-5p* | Rate sensitivity | 5,24E-04 | 4,21E-01 |
| *hsa-miR-301a-3p* | Rate sensitivity | 4,60E-03 | 3,58E-01 |
| *hsa-miR-30c-5p* | Mean insulin | 6,40E-03 | 3,40E-01 |
| | Glucose sensitivity | 3,20E-03 | 3,72E-01 |
| *hsa-miR-30d-5p* | Mean Glucose | 3,13E-04 | -4,46E-01 |
| | Glucose sensitivity | 7,68E-04 | 4,19E-01 |
| | Basal Glucose | 2,84E-03 | -3,76E-01 |
| *hsa-miR-335-5p* | Rate sensitivity | 9,80E-03 | 3,23E-01 |
| | Mean Glucose | 6,87E-04 | -4,20E-01 |
| | Basal Glucose | 2,81E-03 | -3,70E-01 |
| *hsa-miR-340-5p* | Rate sensitivity | 6,62E-03 | 3,39E-01 |
| *hsa-miR-3605-3p* | Rate sensitivity | 9,88E-04 | -4,15E-01 |
| | Glucose sensitivity | 4,87E-03 | -3,53E-01 |
| *hsa-miR-374b-5p* | Rate sensitivity | 7,46E-03 | 3,39E-01 |
| *hsa-miR-4732-3p* | Rate sensitivity | 4,81E-03 | -3,51E-01 |
| *hsa-miR-486-3p* | Rate sensitivity | 1,33E-03 | -3,96E-01 |
| *hsa-miR-6734-5p* | Rate sensitivity | 8,72E-04 | -4,12E-01 |
| | Mean glucose | 5,69E-03 | 3,47E-01 |

| | | | |
|---|---|---|---|
| *hsa-miR-93-3p* | Rate sensitivity | 4,16E-03 | -3,59E-01 |
| *hsa-miR-942-5p* | Glucose sensitivity | 2,55E-03 | -3,74E-01 |
| | Mean glucose | 1,27E-03 | 3,97E-01 |

**Supplementary Table 3.** Selection of n=19 miRNAs for validation through ddPCR technique. Average expression from sequencing, differential expression among groups, and association with Glucose and/or Rate sensitivity are also indicated.

| miRNA | Expression level (Average UMI counts) | Differential Expression among groups | Association with GS | Association with RS |
|---|---|---|---|---|
| hsa-let-7b-5p | 213413,58 | | | X |
| hsa-miR-26a-5p | 19593,70 | | | X |
| hsa-miR-30e-5p | 11540,39 | X | | |
| hsa-miR-30d-5p | 9859,07 | X | X | |
| hsa-let-7c-5p | 2324,78 | | X | |
| hsa-miR-140-3p | 2050,40 | | X | |
| hsa-miR-486-3p | 818,80 | | | X |
| hsa-miR-23b-3p | 700,60 | | X | |
| hsa-miR-374a-5p | 319,42 | X | | |
| hsa-miR-340-5p | 317,60 | X | | X |
| hsa-miR-4732-3p | 312,79 | | | X |
| hsa-miR-335-5p | 310,05 | | | X |
| hsa-miR-942-5p | 300,60 | | X | |
| hsa-miR-34a-5p | 228,04 | X | | |
| hsa-miR-30c-5p | 218,45 | X | X | |
| hsa-miR-2110 | 114,90 | | | X |
| hsa-miR-301a-3p | 110,75 | X | | X |
| hsa-miR-93-3p | 102,63 | | | X |
| hsa-miR-1306-5p | 101,97 | | X | |

**ESM Figure 1**

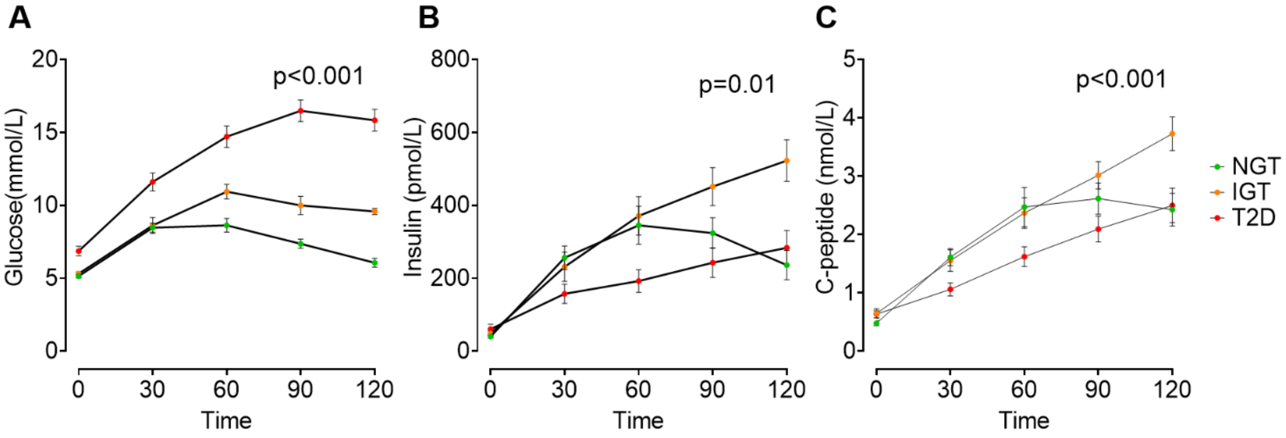


**Supplementary Figure 1. OGTT curves from NGT, IGT and T2D individuals from the discovery cohort.** (**A**) Glucose, (**B**) insulin, (**C**) C-peptide levels during OGTT in NGT (green), IGT (orange) and T2D (red). *p≤0.05, was considered statistically significant for glucose, insulin and C-peptide levels for each time point.

### ESM Figure 2

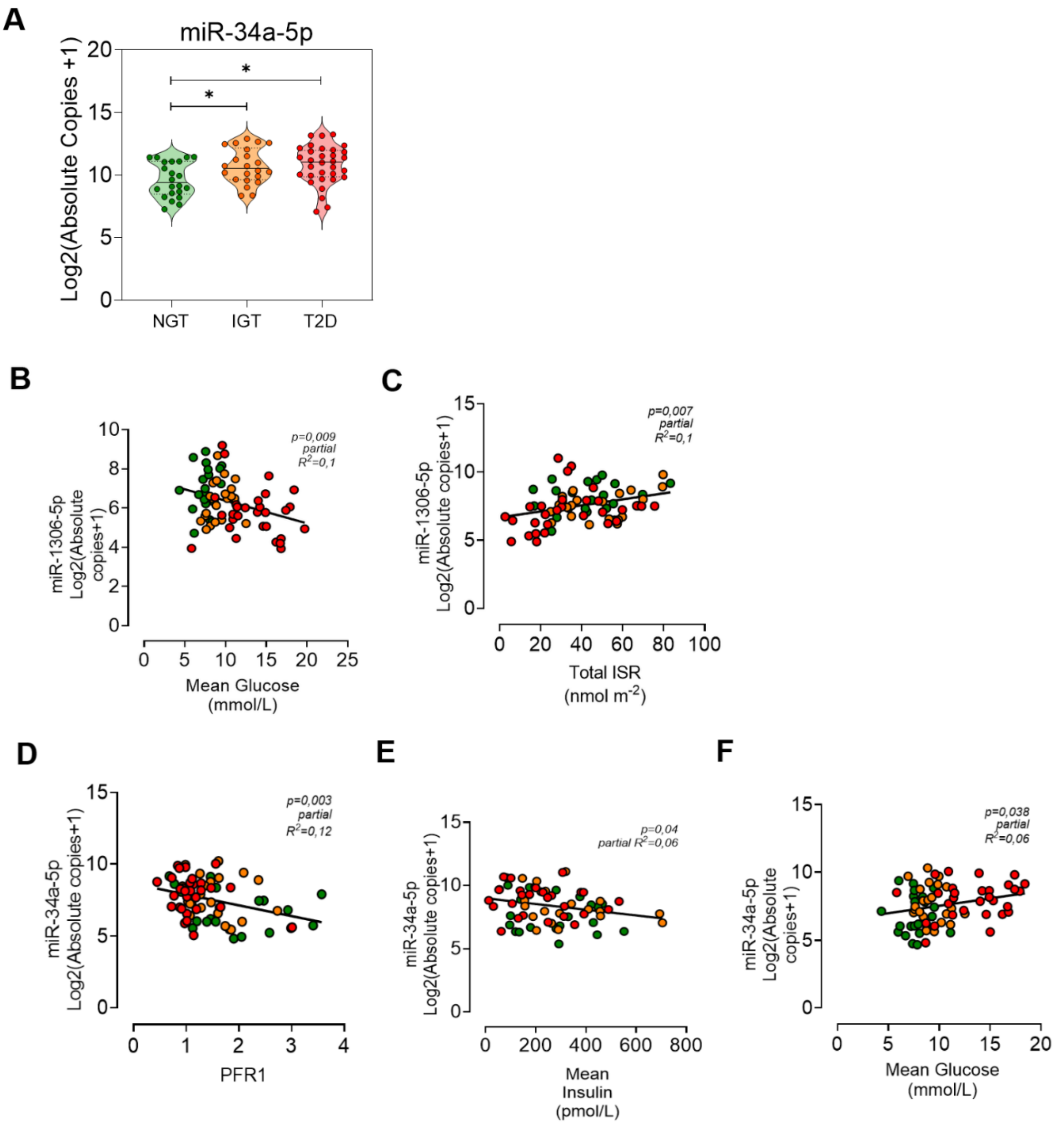


**Supplementary Figure 2. A.** miR-34a expression in NGT, IGT and T2D individuals. Values are presented as Log2 absolute copies. Statistical analysis was performed using the Wald test (DESeq2) with FDR-adjusted $P \leq 0.01$. **B-F.** Scatterplots of covariate-adjusted $log_2$(ddPCR copies + 1) versus beta cell function parameters: **B.** miR-1306-5p vs. mean glucose (mmol/L); **C** miR-1306-5p vs. Total ISR (nmol $m^{-2}$). **D.** miR-34a-5p vs PFR1. **E.** miR-34a-5p vs mean insulin (pmol/L). **F.** miR-34a-5p vs. mean glucose (mmol/L). Data points represent individual donors (NGT: green; IGT: orange; T2D: red) with fitted linear regression.

**ESM Figure 3**

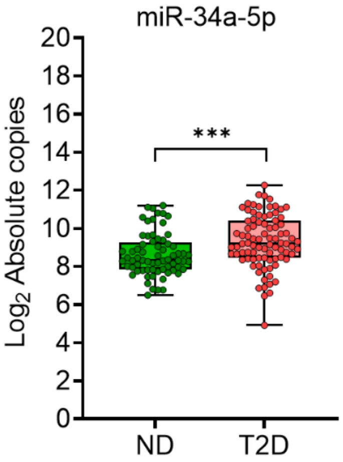


**Supplementary Figure 3.** miR-34a expression in ND and T2D individuals in the validation cohort. Values are presented as log2 absolute copies. Statistical analysis was performed using Wilcoxon test.
